# Systemic cytokines and GlycA discriminate inflammaging, disease progression and corticosteroid response in HTLV-1-associated neuroinflammation

**DOI:** 10.1101/2021.03.16.21253707

**Authors:** Tatiane Assone, Soraya Maria Menezes, Fernanda de Toledo Gonçalves, Victor Angelo Folgosi, Gabriela da Silva Prates, Tim Dierckx, Marcos Braz, Jerusa Smid, Michel E. Haziot, Rosa M N Marcusso, Flávia E. Dahy, Roberta Bruhn, Edward L. Murphy, Augusto César Penalva de Oliveira, Dirk Daelemans, Jurgen Vercauteren, Jorge Casseb, Johan Van Weyenbergh

## Abstract

**Background:** HTLV-1-Associated Myelopathy/Tropical Spastic Paraparesis (HAM/TSP) is an incapacitating neuroinflammatory disorder for which no disease-modifying therapy is available, but corticosteroids provide some clinical benefit.

**Objective:** To investigate systemic cytokines and GlycA as possible biomarkers of immunopathogenesis and therapeutic response to corticosteroid pulse therapy in HAM/TSP.

**Methods:** We prospectively followed 110 People living with HTLV-1 (PLwHTLV-1, 67 asymptomatic individuals and 43 HAM/TSP patients), for a total of 906 person-years. Plasma cytokine levels (IL-2/4/6/10/17A, IFN-γ, TNF) and GlycA were quantified by Cytometric Bead Array and 1NMR, respectively. Cytokine signaling and prednisolone response were validated in an independent cohort by nCounter digital transcriptomics. We applied logistic regression and machine learning algorithms to predict disease progression and glucocorticoid response.

**Results:** IL-6 was positively correlated with age and GlycA in asymptomatics but not HAM/TSP patients. Systemic IFN-γ and IL-17A levels were increased in HAM/TSP patients, as compared to asymptomatics. All patients significantly decreased IL-17A levels post-treatment but only prednisolone-responders decreased IFN-γ levels post-treatment. Higher pre-treatment GlycA and TNF levels significantly predicted a negative therapeutic outcome, which was associated with higher post-treatment IFN-γ levels. Low IL-4 and IL-10 levels in incident HAM/TSP can be reverted to increased IL-10 and IL-4/IL-13 signaling by prednisolone in vitro.

**Conclusions:** 1) An age-related increase in systemic IL-6/GlycA levels reveals inflammaging in PLwHTLV-1. 2) IFN-γ and IL-17A are biomarkers of untreated, active HAM/TSP disease, while pre-treatment GlycA and TNF predict therapeutic response to prednisolone pulse therapy. 3) Low IL-4/IL-10 and high IFN-γ signaling in incident HAM/TSP can be normalized by prednisolone.

## INTRODUCTION

Human T-cell Lymphotropic Virus type-1 (HTLV-1) is unique as it is both oncogenic[1,2] and capable of triggering HTLV-1-Associated Myelopathy/Tropical Spastic Paraparesis (HAM/TSP) and other inflammatory diseases[3-5]. Worldwide, 10 million people are estimated to be living with HTLV-1 (PLwHTLV-1)[3], of which 1-2% develop HAM/TSP, an incapacitating neuroinflammatory disorder with similarity to primary progressive multiple sclerosis[4,5]. Currently, no disease-modifying therapy is available for HAM/TSP but corticosteroids and other immunomodulators (IFN-α, cyclosporin) provide some clinical benefit[5,6]. Moreover, we recently demonstrated HAM/TSP is an independent predictor for early mortality among PLwHTLV-1[7]. As such, biomarkers to predict and/or monitor disease progression for PLwHTLV-1 and therapeutic outcome in HAM/TSP patients are direly needed. Acute or chronic inflammation, measured by systemic cytokines or glycoprotein acetylation (GlycA) respectively, can reliably predict long-term outcomes of inflammatory and infectious diseases in large prospective population studies[8,9], but have not been explored in combination in HTLV-1 infection or HAM/TSP.

## RESULTS AND DISCUSSION

As shown in Table 1, HAM/TSP patients were age- and gender-matched to HTLV-1-infected controls without neurological symptoms (asymptomatics, AS), while proviral load was increased in HAM/TSP, as expected[4,5]. Since age and gender are major determinants of HAM/TSP pathogenesis, as disease onset usually occurs after several decades and women are more affected[4,5], we investigated if cytokines or GlycA were linked to demographics. Gender did not influence systemic cytokine levels (not shown), whereas both IL-6 (Spearman’s ρ=0.36, p=0.00018) and IL-10 (ρ=0.22, p=0.021) were positively correlated with age in PLwHTLV-1 (Fig. 1A-1D). Surprisingly, this age-dependent cytokine increase was specific to AS (Fig. 1B-1E), and absent in HAM/TSP (Fig. 1C-1F). In contrast, chronic inflammation marker GlycA did not correlate with age (p=0.12), but was higher in females (p=0.0069, Fig. 1G). Among all cytokines, only IL-6 was significantly correlated to GlycA in AS (Fig. 1H, p=0.00049, ρ=0.45) but not HAM/TSP patients (p=0.16). Among pro-inflammatory cytokines, IL-6 uniquely predicts global functional decline in aging[10] and inflammaging in a systematic review and meta-analysis[11]. Lifelong chronic infection with other latent viruses (CMV and HIV) causes long-term activation of the immune system over time, contributing to inflammaging[12]. To our knowledge, this study is the first demonstration of inflammaging in PLwHTLV-1, which also corroborates the increased mortality rate we observed in the same cohort[7]. Of interest, none of the cytokines nor GlycA were significantly correlated to proviral load (Fig. 1I and not shown), but a tendency was observed for IFN-γ (Fig. 1I, ρ=0.20, p=0.054).

**Table 1.**
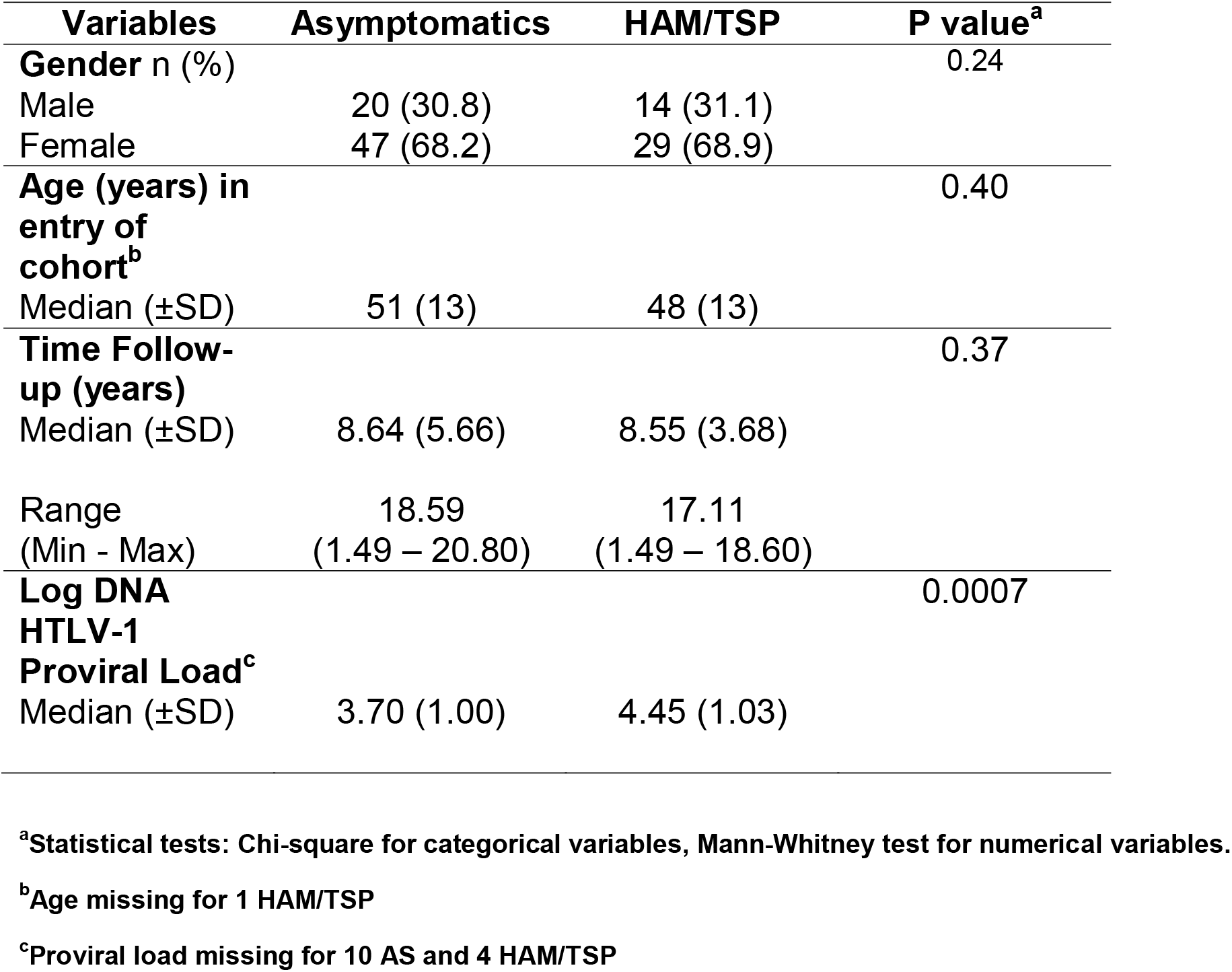
Principal characteristics of the participants (AS vs. HAM/TSP)

**Legend Fig. 1:**
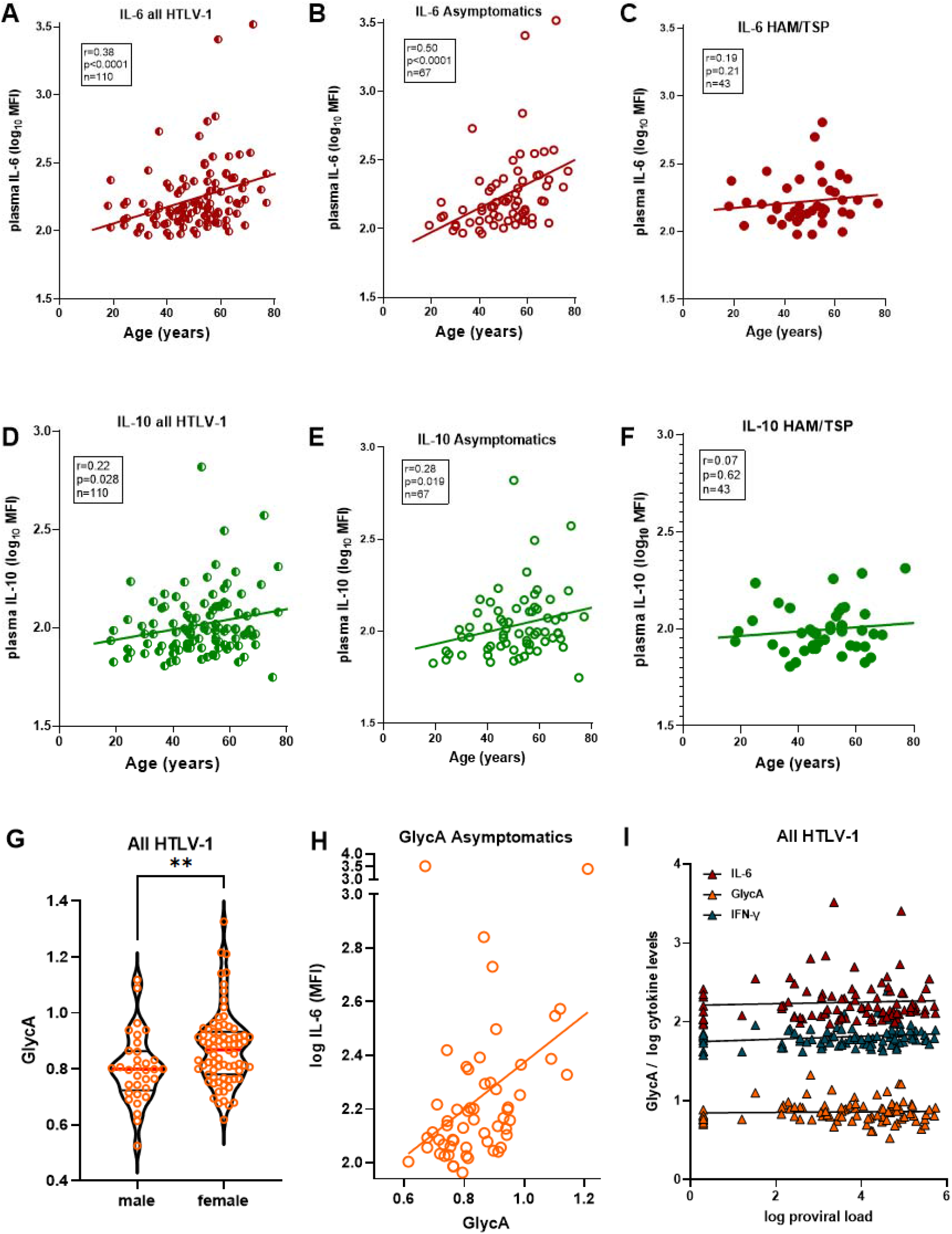
Age and gender differentially affect cytokines and chronic inflammation marker GlycA in people living with HTLV-1, independent of proviral load. (A) IL-6 levels are significantly correlated to age at sampling in all PLwHTLV-1, which is driven by the strong correlation in asymptomatic individuals (AS) (B), which is absent in HAM/TSP patients (C). (D) IL-10 levels are significantly correlated to age at sampling in all PLwHTLV-1, which is driven by the strong correlation in AS (E), which is absent in HAM/TSP patients (F). (G) GlycA levels are significantly higher in female PLwHTLV-1 (p=0.0069, Mann-Whitney test). (H) GlycA levels are positively correlated to IL-6 levels in AS only (p=0.00049, ρ=0.45). (I) Proviral load is not significantly correlated to IL-6 (ρ=0.07, p=0.45) or GlycA levels (ρ=-0.06, p=0.72) in PLwHTLV-1, while a tendency is observed for IFN-γ (r=0.20, p=0.054). Correlation is determined using Spearman’s method, uncorrected p-values are reported, significant correlations of IL-6 with age and with GlycA were robust to correction for multiple testing (Bonferroni p<0.05).

When comparing cytokine levels between clinical groups, we observed a significant increase in IFN-γ (p=0.007) and IL-17A (p=0.0001) in HAM/TSP patients, as compared to AS (Fig. 2A), while other cytokines and GlycA did not differ (not shown). Using logistic regression (detailed in Suppl. Table), we found that IL-17A and proviral load were independently associated with clinical status, consistent with a weak correlation between IFN-γ and proviral load (Fig. 1I). However, logistic regression resulted in low classification accuracy for HAM/TSP patients, as only 25/39 (64.1%) were correctly classified, in contrast to 49/57 (86.0%) of correctly predicted AS (ROC AUC 0.85[0.77-0.93]). Therefore, we used machine learning algorithms to improve classification, which revealed a decision tree classifying 39/43 HAM/TSP and 58/67 AS, respectively with 90.7% and 86.6% accuracy (ROC AUC 0.87, Fig. 2B). Among the first leaves in this decision tree are IL-17A and IL-10, confirming previous findings in a UK cohort[13]. In addition, we used Bayesian network learning to identify direct vs. indirect associations between cytokines, GlycA, clinical and demographic data[14]. As shown in Fig. 2C, only IL-17A was directly connected to clinical status, while all other cytokines were ‘upstream’ of IL-17A. This Bayesian network revealed a direct link between GlycA and IL-6, whereas the observed correlation between age and IL-6 (Fig. 1A-B) appears dependent on TNF and IFN-γ, the latter directly influencing GlycA. IFN-γ was found upstream of all other cytokines and, consequently, of disease status, which underscores the previously identified IFN gene signature in HAM/TSP [15]. In this unsupervised model, proviral load was not significantly associated with the other parameters and hence absent from the network.

**Legend Fig. 2:**
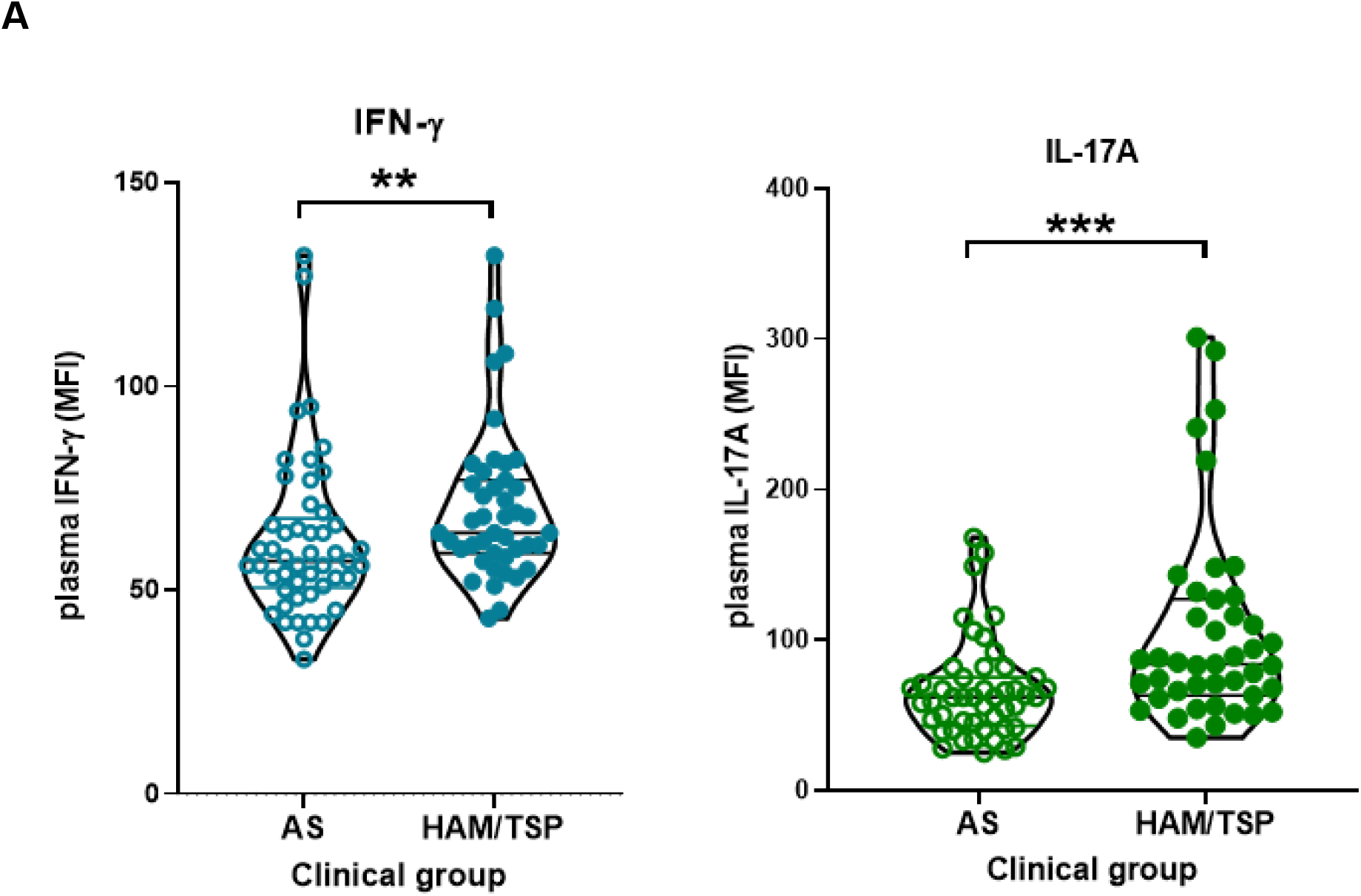
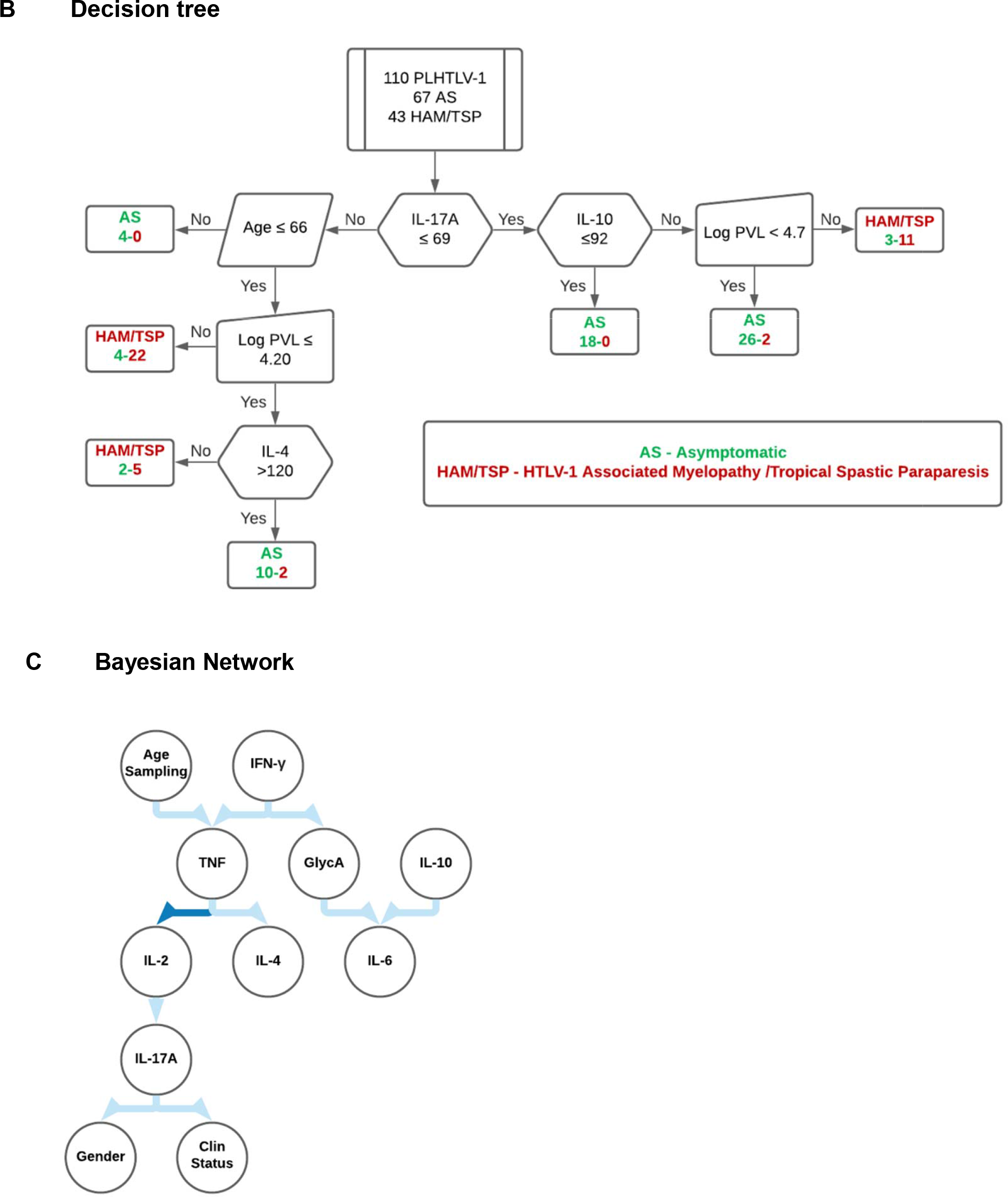
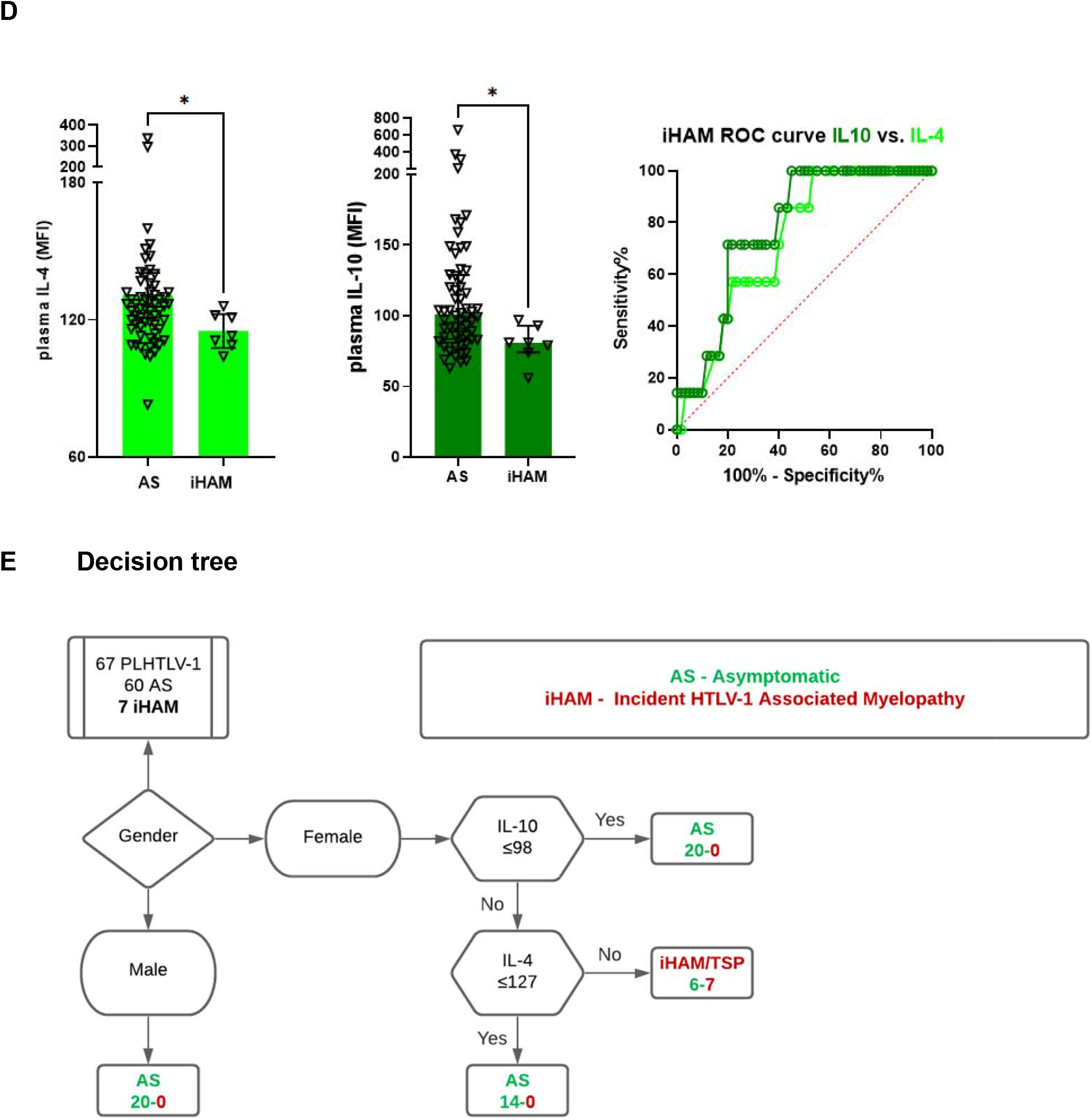
Active HAM/TSP disease status is characterized by increased IFN-γ and IL-17A, whereas a decrease in anti-inflammatory cytokines IL-4 and IL-10 predicts incident disease (iHAM) during follow-up. (A) Among all cytokines tested, only IFN-γ (p=0.0054) and IL-17A (p<0.0001) levels in HAM/TSP patients (n=43) were significantly different (Mann-Whitney test), as compared to asymptomatic individuals (AS, n=67). (B) Machine learning-derived decision tree discriminating AS from HAM/TSP patients (J48 pruned tree) with 90% accuracy (97 out of 110 PLwHTLV-1 correctly classified, of which 61/67 AS and 38/43 HAM/TSP patients, ROC AUC 0.97, Kappa statistic 0.79). (C) Bayesian Network representing the strongest associations between cytokines, GlycA, clinical and demographic data (strength of arcs as defined previously[14]: dark blue 100x, light blue 10x). Proviral load was not significantly associated with the other parameters and hence not shown. (D) Lower IL-4 (left panel) and IL-10 (middle panel) at the first sample (at entry in the cohort) significantly predicted incident HAM/TSP (iHAM), as compared to AS who remained neurologically asymptomatic during follow-up (right panel, ROC AUC 0.78 and 0.73, respectively). (E) Machine learning revealed a decision tree discriminating AS from incident HAM/TSP (PART decision tree) with 91.0% accuracy (54/60 AS and 7/7 iHAM, ROC AUC 0.94).

During 906 person-years of clinical follow-up, seven incident cases of HAM/TSP were observed, all women, in agreement with a higher risk of clinical progression in female PLwHTLV-1. Lower IL-10 and IL-4 levels (Mann-Whitney p=0.013 and p=0.043, respectively, Fig. 2D) at entry in the cohort significantly predicted clinical progression to definite HAM/TSP, as compared to AS who remained neurologically asymptomatic during follow-up, as confirmed by univariate logistic regression (IL-10: AUC 0.78[0.65-0.92]; IL-4: AUC 0.73[0.58-0.88]). Multivariate logistic regression was not possible due to perfect separation (gender), but similar results were obtained if only female AS were used for comparison to incident HAM/TSP. Machine learning generated a decision tree discriminating AS from incident HAM/TSP with 91.0% accuracy (54/60 AS and 7/7 iHAM, ROC AUC 0.95, Fig. 2E).

Next, we investigated if cytokines and GlycA might be candidate biomarkers for therapeutic response in HAM/TSP patients. In this cohort, all eligible patients were uniformly treated with intravenous methylprednisolone pulse therapy, which allowed comparisons before and after treatment. Patients with >1 year pulse therapy follow-up were classified as responders (n=13) and non-responders (n=25), based on changes in Osame Motor Disability Score (decrease or stable: responders, increase: non-responders). All patients significantly decreased IL-17A levels after treatment (Fig. 3A, p=0.013), while strong variability but no directionality was observed for any other cytokine, nor for GlycA (Fig. 3A). However, only prednisolone-responders significantly decreased IFN-γ levels after treatment (p=0.008). In addition, pre-treatment TNF levels were significantly associated with therapeutic outcome (Fig. 3B, p=0.037). In addition, pre-treatment GlycA levels were able to predict therapeutic response, as measured by quantitative changes in Osame Motor Disability Score, either by itself (Fig. 3C, left panel) or as a combined TNF/GlycA score (Fig. 3C, right panel). This finding, in addition to GlycA’s relative stability over time[9], underscores the potential of GlycA as a clinically useful biomarker in HAM/TSP.

**Legend Fig. 3:**
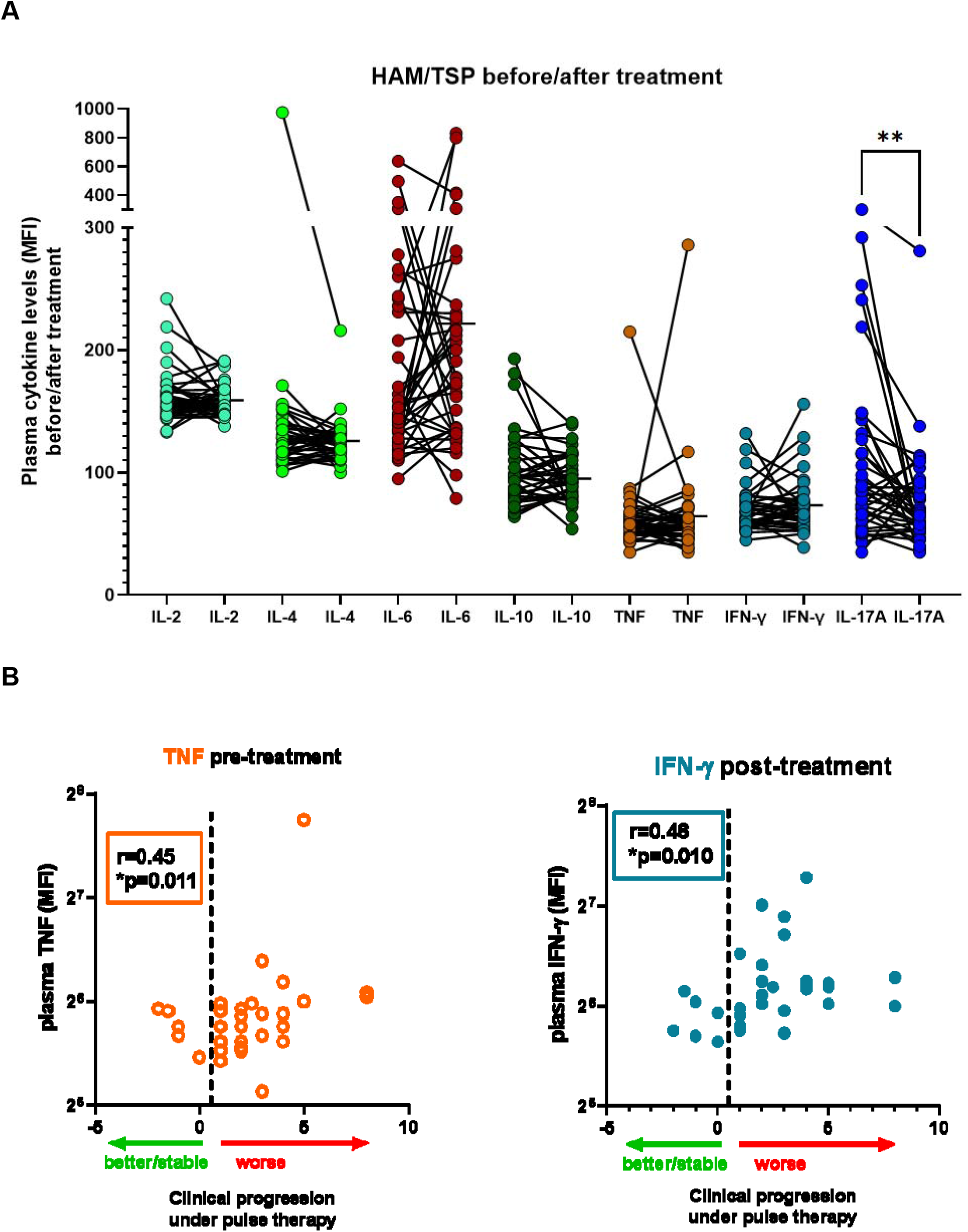
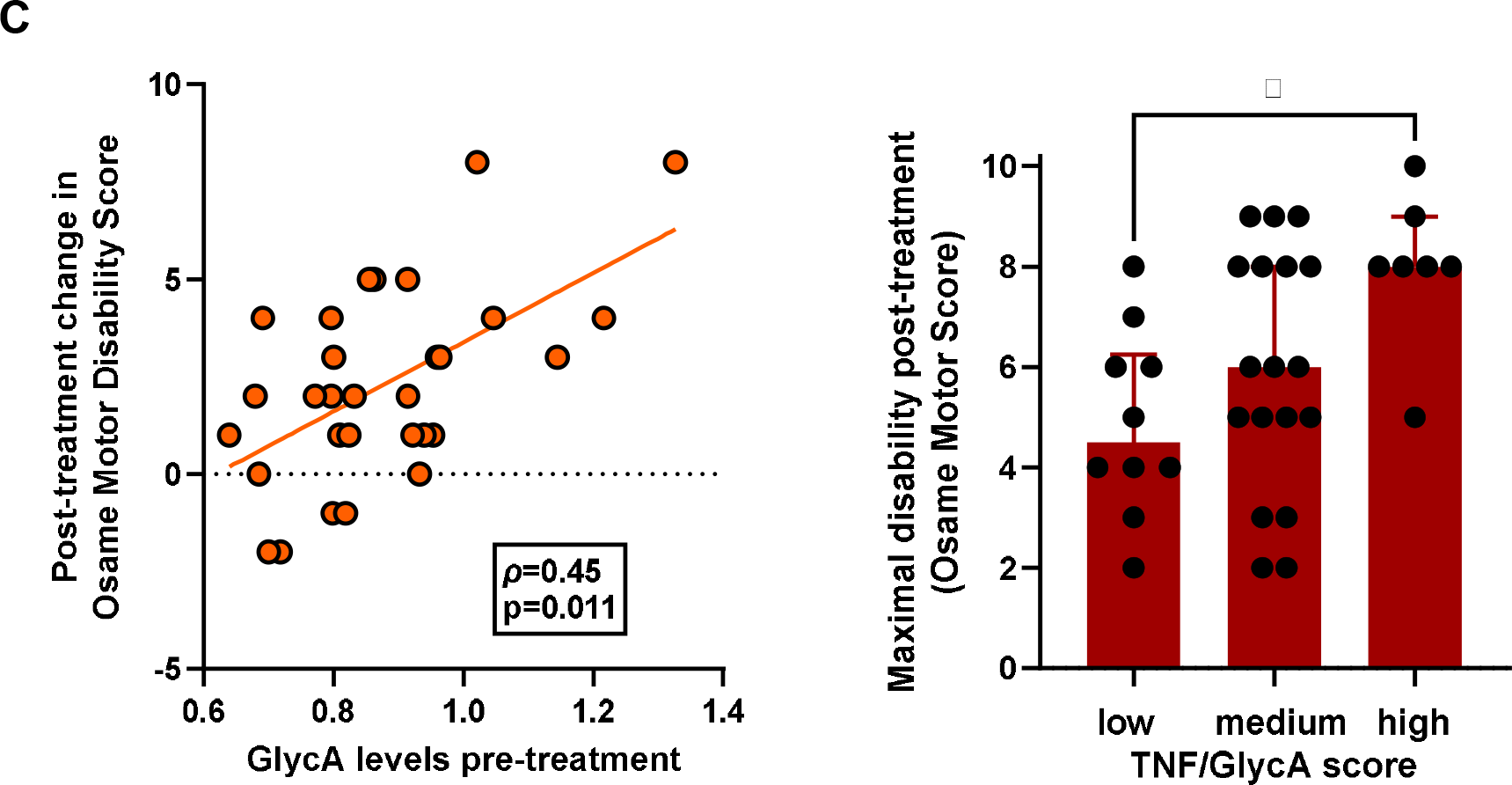
Cytokines and GlycA predict therapeutic success vs. failure of methylprednisolone pulse therapy in HAM/TSP patients. (A) Among all plasma cytokines measured in HAM/TSP patients, only IL-17A significantly decreases after pulse therapy with intravenous methylprednisolone (Wilcoxon test p=0.003). (B) Pre-treatment TNF (left panel) and post-treatment IFN-γ (right panel) are correlated with the magnitude of clinical worsening after prednisolone pulse therapy (Spearman correlation). (C) Pre-treatment GlycA levels are correlated with the magnitude of clinical worsening after methylprednisolone pulse therapy (Spearman correlation, left panel). A higher TNF/GlycA score (0.025*TNF + 9.18*GlycA - 7.28) predicts worse Osame Motor Disability Score after methylprednisolone pulse therapy (right panel, ANOVA with Bonferroni post-test, *p=0.010).

Finally, we used digital transcriptomics (nCounter) to provide broader mechanistic insight into the cytokine signaling pathways mediating HAM/TSP disease progression[16,17], and the effect of prednisolone treatment *in vitro*. From a well-characterized US cohort of PLwHTLV-1[18], we selected 4 AS and 4 HAM/TSP patients(including the only two incident HAM/TSP cases from the entire HTLV-1 cohort), and age-, gender- and ethnicity-matched healthy controls (n=4). First, we confirm that exacerbated IFN signaling is a hallmark of HAM/TSP disease progression[15], being significantly higher in both stable and incident HAM/TSP (Fig. 4A), as compared to healthy controls and AS. The IFN signaling pathway was effectively and homogeneously down-regulated in all clinical groups by prednisolone treatment *in vitro* (Fig. 4A), also reflected by decreased expression of the IFN-regulated MHC Class I antigen presentation pathway (Fig. 4A). In contrast, *in vitro* prednisolone treatment did not have a uniform effect on IL-10 signaling (Fig. 4B), IL4/IL-13 signaling (Fig. 4C) or IL-2 cytokine family signaling (Fig. 4D). We found that prednisolone stimulation of the IL-10 and IL-4/IL-13 signaling pathways were strongly influenced by clinical status, increasing in HAM/TSP and highest in the two patients with incident HAM/TSP (Fig. 4B-4C, Kruskal-Wallis, Dunn’s post-test, p<0.05). Therefore, the defective anti-inflammatory IL-4 and IL-10 production we observed in the incident HAM/TSP cases (at the protein level, Fig. 2C), might be effectively restored by early prednisolone therapy. Recent clinical guidelines for HAM/TSP suggest that incident/early HAM/TSP patients might benefit most from corticosteroid therapy[6]. Supporting this hypothesis, two of the incident HAM/TSP cases from our Brazilian cohort have completed >1 year of pulse therapy and were indeed classified as “responders” (stable Osame Motor Disability). As shown in Fig. 4E, IL-17 signaling is strongly increased by prednisolone treatment *in vitro*, across all clinical groups, in strong contrast to the decreased systemic IL-17A levels we observed in HAM/TSP patients after *in vivo* treatment. This apparent contradiction coincides, however, with demonstrated ex vivo (increased IL-17A plasma levels[13]) vs. in vitro differences (decreased Th17/intracellular IL-17A[19,20]) in HAM/TSP, which might indicate cell type-specific effects of prednisolone pulse therapy *in vivo*, considering that the cellular source of plasma IL-17A is unknown. Likewise, IL17A levels were not correlated with CD4+ levels quantified by flow cytometry at baseline (not shown). Noteworthy, neutrophils have recently been identified as microbiome-driven IL-17 producers[21], opening potentially new research avenues in HTLV-1 pathogenesis.

**Legend Fig. 4:**
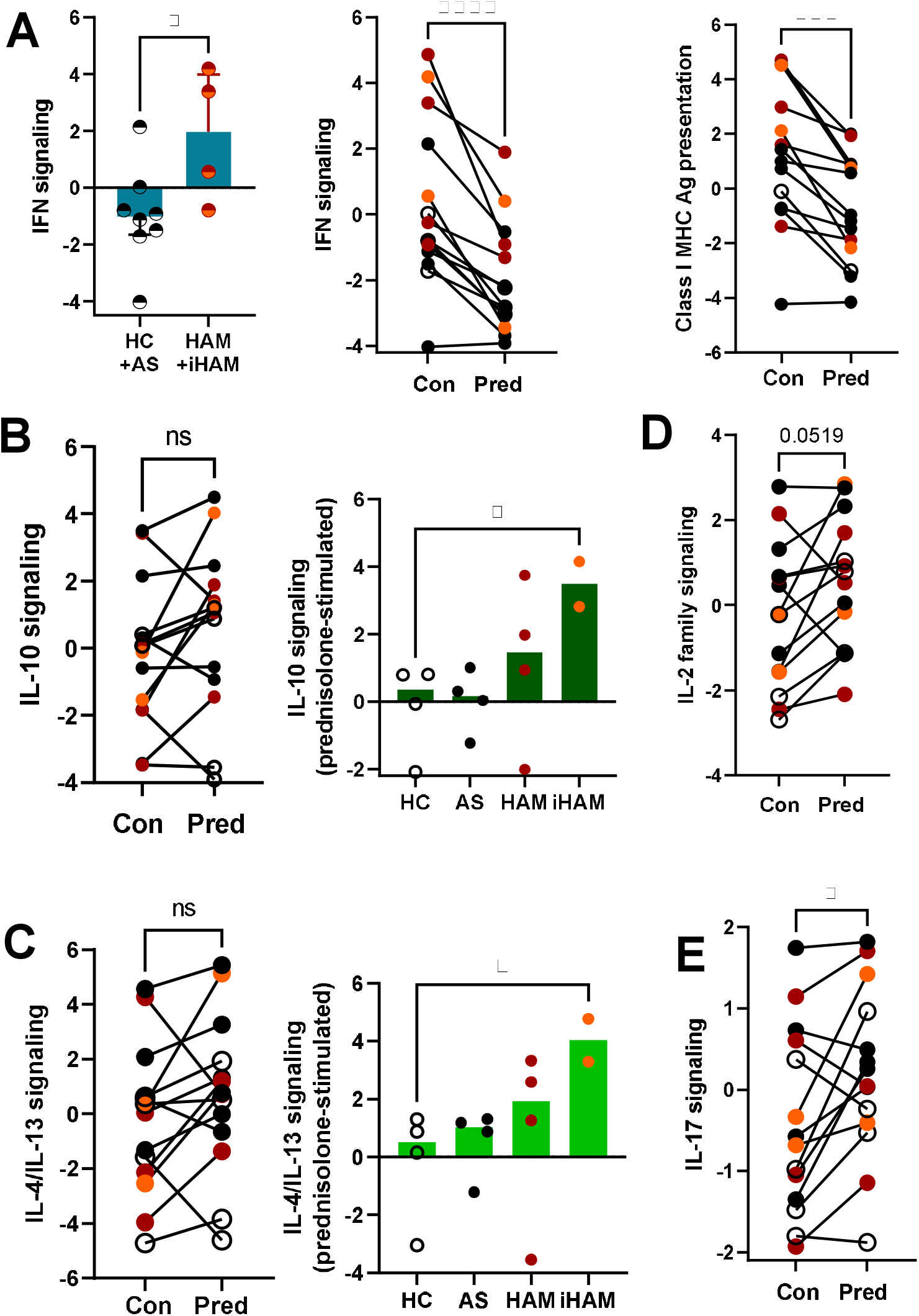
Transcriptomic validation of cytokine signaling pathways and prednisolone response in an independent cohort of PLwHTLV-1. Digital transcriptomics (nCounter) was used to quantify cytokine signaling pathways inHAM/TSP disease progression, and the effect of in vitro prednisolone treatment in PBMCs from four AS, four HAM/TSP patients (including two incident HAM/TSP), and four age-, gender- and ethnicity-matched healthy controls. (A). IFN signaling is significantly higher in both stable and incident HAM/TSP (left panel), as compared to age-, gender-, and ethnicity-matched healthy controls and AS. IFN signaling was homogeneously down-regulated in all clinical groups by prednisolone treatment in vitro (middle panel, Wilcoxon test p<0.0001). Down-regulation was confirmed by decreased expression of the IFN-regulated MHC Class I antigen presentation pathway (right panel, Wilcoxon test p<0.001). (B) In vitro prednisolone treatment does not have a uniform effect on IL-10 signaling (left panel), but depends on clinical status, significantly higher in incident HAM/TSP (right panel, Kruskal-Wallis with Dunn’s post-test, p<0.05). (C) In vitro prednisolone treatment does not have a uniform effect on IL-4/IL-13 signaling (left panel), but depends on clinical status, highest in incident HAM/TSP (right panel, Kruskal-Wallis with Dunn’s post-test, p<0.05). (D) In vitro prednisolone treatment does not significantly impact IL-2 cytokine family signaling. (E) In vitro prednisolone treatment significantly increases IL-17 signaling, independent of clinical groups. HC: Healthy controls (open circles); AS: Asymptomatics (black circles); iHAM: incident HAM/TSP (orange circles); stable HAM/TSP (red circles); Con: untreated in vitro PBMCs; Pred: Prednisolone-treated PBMCs in vitro.

In contrast to IL-17A, exacerbated IFN-γ production (ex vivo and in vitro) has been demonstrated by several groups, including ours, as a hallmark of HAM/TSP[20,22-25]. However, this is the first study to show that plasma IFN-γ and IL-17A increase in untreated, active disease and are differentially impacted by corticosteroid therapy: IFN-γ levels decrease in responders only, while IL-17A levels decrease uniformly for all patients.

In conclusion, we found that decreased anti-inflammatory cytokines IL-4 and IL-10 are major risk factors for incident HAM/TSP in PLwHTLV-1. On the other hand, GlycA and inflammatory cytokines IL-6, TNF and IFN-γ are promising candidate biomarkers for immunomonitoring of inflammaging in PLwHTLV-1, and of disease progression and corticosteroid therapeutic response in HAM/TSP patients. In addition, we provide predictive regression models and decision trees, which could be prospectively tested in clinical trials or independent cohort studies.

## Data Availability

All data are available in the manuscript or in the Supplementary data.

## Acknowledgments

We would like to thank all the interns of the IIER Neurology program, the patients and their relatives for their participation.

## Notes

### Competing Interest Statement

The authors have declared no competing interest.

### Funding Statement

This research was funded by FAPESP grant numbers 2017/08320-5, 2018/07239-2 and 2016/03025-2 (scholarship to TA); CNPq (Scholarship to JC), FFM (support to JC), FWO (grant G0A0621N to JVW) and KU Leuven (Vaast Leysen Leerstoel voor Infectieziekten in Ontwikkelingslanden, to JVW).

### Author Declarations

The Ethical Board of the IIER (Instituto de Infectologia Emilio Ribas, Sao Paulo, Brazil) approved the protocol (Number 07688818.2.1001.0061). Signed informed consent was obtained from all participants prior to study inclusion.

