## Supplementary Methods & Suppl. Table for "Systemic cytokines and GlycA discriminate inflammaging, disease progression and corticosteroid response in HTLV-1-associated neuroinflammation"

**Cohort characteristics**

The open cohort investigated in this study was set up in 1997 and currently has >1100 PLwHTLV-1 in regular follow-up, with new patients added at a rate of approximately 50 per year. All volunteers underwent serological screening for HTLV-1 at the “Emilio Ribas” Institute of Infectious Diseases, using GOLD ELISA HTLV-1/ 2 (Diasorin, UK), followed by confirmation with Western Blot (MP Diagnostics, HTLV Blot 2.4®) and in-house nested PCR [1].

Blood samples were collected in K_3_-EDTA (0.054 ml/tube), plasma was separated by centrifugation (15 min, 2500 rpm) and PBMC were purified by Ficoll density gradient centrifugation (GE Healthcare Life, USA ). Cells were washed with saline solution; cell number was adjusted to 106 cells and then stored at -80°C. DNA was extracted using a commercial kit (Illustra Tissue and Cells Genomic Prep Mini Spin kit, Easton Turnpike, Fairfield, CA) according to the manufacturer’s instructions, and stored at -80 °C for later analysis.

**HTLV-1 proviral load (PVL)**

HTLV-1 proviral load was quantified by real-time PCR, using primers and probes targeting the pol gene: SK110 and SK111, albumin gene: AlbS and AlbAS, the internal HTLV-1 Taq Man probe was selected using Oligo (National Biosciences). All samples were performed in duplicate, and results expressed as HTLV-1 DNA copies/10^4^ PBMCs, as described elsewhere [1].

**Neurological evaluation and HAM/TSP diagnosis**

PLwHTLV-1 were classified in two groups according to their neurological status: 65 asymptomatic HTLV-1-infected individuals and 45 HAM/TSP patients, of which 38 were followed up after treatment with methylprednisolone (1g intravenously, every 45 days). HAM/TSP diagnostic criteria was based on recommendations from an international consortium [2]. Clinical evaluation and a standardized screening neurological examination were performed by a board-certified neurologist, blinded for HTLV-1 clinical status for all subjects.

**Plasma cytokine levels**

Concentrations of plasma cytokines were measured using the CBA (Cytometric Bead Array, BD Biosciences) Human Th1/Th2/Th17 Cytokine Kit, including Interleukin-2 (IL-2), Interleukin-4 (IL-4), Interleukin-6 (IL-6), Interleukin-10 (IL-10), Tumor Necrosis Factor (TNF), Interferon-γ (IFN-γ), and Interleukin-17A (IL-17A), in accordance with the manufacturer’s instructions.

**GlycA (Glycoprotein Acetyl) quantification in plasma**

GlycA concentration was quantified using the Nightingale Health Ltd. high-throughput metabolomics platform (Helsinki, Finland) [3]. Briefly, a ^1^H-NMR spectrum is taken from 350 μl of plasma, with the area under the peak measured at approximately 2 ppm quantifying signal originating from N-acetyl sugar groups present on acute phase glycoproteins (α-1-acid glycoprotein, α-1-antitrypsin, α-1-antichymotryspin, haptoglobin, transferrin).

**Data Quality Control**

Data collection was performed by the first author with the help of neurologists and infectologists of the HTLV outpatient clinic at the “Emilio Ribas” Institute for Infectious Diseases. Data entry in the electronic database RedCap [4] was performed by two administrative assistants, and subsequently checked by the first and last author.

**Ethical Issues**

The Ethical Board of the IIER (“Instituto de Infectologia Emilio Ribas”, Sao Paulo, Brazil) approved the protocol (Number 07688818.2.1001.0061), signed informed consent was obtained from all participants prior to study inclusion.

**Statistical analysis, Machine Learning and Bayesian Network analysis**

Statistical analysis was performed using XLStat and GraphPad Prism version 9, San Diego, CA). Logistic regression and non-parametric statistical tests (Mann–Whitney, Wilcoxon tests, Kruskall-Wallis and Spearman correlation) were used, except for Maximal Osame Motor Disability Score (which followed normal distribution, ANOVA), with Bonferroni correction for multiple comparisons as indicated in the text. Machine learning algorithms (attribute selection, J48 and PART decision trees) were performed using Weka (version 3.8.4). Bayesian network analysis was performed as previously described [5].

### Digital transcriptomics and biological pathway analysis

Digital transcriptomic analysis (nCounter, Nanostring Technologies) and biological pathway analysis of *in vitro* prednisolone response was performed as previously described [6,7] , using the Myeloid/Innate Immunity Panel, consisting of >700 host genes, as well as customized HTLV-1 Hbz and Tax probes. PBMCs were obtained from an independent, previously characterized cohort of PLwHTLV-1 (4 asymptomatic, 4 HAM/TSP patients), as well as age-, gender- and ethnicity-matched healthy controls (n=4) from the HOST study [8].

**Supplementary Table: Multivariate logistic regression (Asymptomatics (AS) vs. HAM/TSP patients)**

| **Model 1** |  |  |  |  |
| --- | --- | --- | --- | --- |
| **Parameter estimates** | **Variable** | **Estimate** | **Standard error** | **95% CI (profile likelihood)** |
| β0 | **Intercept** | -5.607 | 1.911 | -9.758 to -2.213 |
| β1 | **Gender** | 0.5821 | 0.5803 | -0.5570 to 1.740 |
| β2 | **Age sampling** | -0.021 | 0.01928 | -0.06009 to 0.01646 |
| β3 | **log PVL** | 0.9479 | 0.2756 | 0.4702 to 1.561 |
| β4 | **IL-17A** | 0.0279 | 0.009137 | 0.01207 to 0.04782 |
| **Odds ratios** | **Variable** | **Estimate** | **95% CI (profile likelihood)** |  |
| β0 | Intercept | 0.003671 | 5.782e-005 to 0.1094 |  |
| β1 | Gender | 1.79 | 0.5729 to 5.699 |  |
| β2 | Age sampling | 0.9792 | 0.9417 to 1.017 |  |
| β3 | log PVL | 2.58 | 1.600 to 4.765 |  |
| β4 | IL-17A | 1.028 | 1.012 to 1.049 |  |
| **Area under the ROC curve** |  |  |  |  |
| Area | 0.8525 |  |  |  |
| Std. Error | 0.04051 |  |  |  |
| 95% confidence interval | 0.7731 to 0.9318 |  |  |  |
| P value | <0.0001 |  |  |  |
| **Classification table** | **Predicted 0** | **Predicted 1** | **Total** | **% Correctly classified** |
| Observed 0 | 51 | 6 | 57 | 89.47 |
| Observed 1 | 12 | 27 | 39 | 69.23 |
| Total | 63 | 33 | 96 | 81.25 |
| **Hypothesis tests** | **Statistic** | **P value** |  |  |
| Log-likelihood ratio (G squared) | 40.11 | <0.0001 |  |  |
| **Model 2** |  |  |  |  |
| **Parameter estimates** | **Variable** | **Estimate** | **Standard error** | **95% CI (profile likelihood)** |
| β0 | **Intercept** | -6.619 | 1.526 | -10.02 to -3.985 |
| β1 | **log PVL** | 0.9814 | 0.2739 | 0.5038 to 1.590 |
| β2 | **IL-17A** | 0.02763 | 0.008742 | 0.01243 to 0.04664 |
| **Odds ratios** | **Variable** | **Estimate** | **95% CI (profile likelihood)** |  |
| β0 | Intercept | 0.001335 | 4.450e-005 to 0.01859 |  |
| β1 | log PVL | 2.668 | 1.655 to 4.903 |  |
| β2 | IL-17A | 1.028 | 1.013 to 1.048 |  |
| **Area under the ROC curve** |  |  |  |  |
| Area | 0.8538 |  |  |  |
| Std. Error | 0.04077 |  |  |  |
| 95% confidence interval | 0.7739 to 0.9337 |  |  |  |
| P value | <0.0001 |  |  |  |
| **Classification table** | **Predicted 0** | **Predicted 1** | **Total** | **% Correctly classified** |
| Observed 0 | 49 | 8 | 57 | 85.96 |
| Observed 1 | 14 | 25 | 39 | 64.1 |
| Total | 63 | 33 | 96 | 77.08 |
| **Hypothesis tests** | **Statistic** | **P value** |  |  |
| Log-likelihood ratio (G squared) | 38.13 | <0.0001 |  |  |
| **Data summary** |  |  |  |  |
| Number of individuals | 110 |  |  |  |
| Missing data (proviral load, PVL) | 14 |  |  |  |
| Rows analyzed (#observations) | 96 |  |  |  |
| Number of HAM/TSP | 39 |  |  |  |
| Number of AS | 57 |  |  |  |
| Number of parameter estimates | 5 |  |  |  |
| #observations/ #parameters | 19.2 |  |  |  |
| # of HAM/TSP/#parameters | 7.8 |  |  |  |
| # of AS/#parameters | 11.4 |  |  |  |
